# The initial impact of a national BNT162b2 mRNA COVID-19 vaccine rollout

**DOI:** 10.1101/2021.04.26.21256087

**Authors:** Ahmed Zaqout, Joanne Daghfal, Israa Alaqad, Saleh A.N. Hussein, Abdullah Aldushain, Muna A. Almaslamani, Mohammed Abukhattab, Ali S. Omrani

## Abstract

**Objective:** We herein report the initial impact of a national BNT162b2 rollout on SARS-CoV-2 infections in Qatar.

**Methods:** We included all individuals who by 16 March 2021 had completed ≥14 days of follow up after the receipt of BNT162b2. We calculated incidence rates (IR) and their 95% confidence intervals (CI), during days 1–7, 8–14, 15–21, 22–28, and >28 days post-vaccination. Poisson regression was used to calculate incidence rate ratios (IRR) relative to the first 7-day post-vaccination period.

**Results:** We included 199,219 individuals with 6,521,124 person-days of follow up. SARS-CoV-2 infection was confirmed in 1,877 (0.9%), of which 489 (26.1%) were asymptomatic and 123 (6.6%) required oxygen support. The median time from first vaccination to SARS-CoV-2 confirmation was 11.9 days (IQR 7.7–18.2). Compared with the first 7-day post-vaccination period, SARS-CoV-2 infections were lower by 65.8–84.7% during days 15–21, days 22–28, and >28 days (P <0.001 for each). For severe COVID-19, the incidence rates were 75.7– 93.3% lower (P <0.001 for each) during the corresponding time periods.

**Conclusion:** Our results are consistent with an early protective effect of BNT162b2 against all degrees of SARS-CoV-2 severity.

## Background

A two-dose regimen of BNT162b2, the Pfizer-BioNTech COVID-19 mRNA vaccine, was shown to reduce the risk of SARS-CoV-2 by around 95% in a randomized clinical trial, and in a mass national vaccination program.[1, 2] On 23 December 2020, Qatar started a national BNT162b2 rollout programme, in addition to existing COVID-19 public health control measures. The rollout initially prioritised healthcare workers, individuals aged ≥50 years, and those with chronic or immune suppressive medical conditions. We herein report the initial impact of BNT162b2 on SARS-CoV-2 infections in Qatar.

## Methods

SARS-CoV-2 infections were confirmed using real-time PCR on upper or lower respiratory samples. BNT162b2 was supplied in multidose 0.45 mL vials, and was stored, prepared and administered according to the manufacturer’s instructions.[3] We used the COVID-19 database at the Communicable Disease Center, Hamad Medical Corporation, to retrospectively collate clinical and outcome data for all individuals who by 16 March 2021 had completed ≥14 days of follow up after the receipt of at least one dose of BNT162b2. Confirmed COVID-19 and severe COVID-19 were defined according to the Food and Drug Administration criteria.[1]

We calculated SARS-CoV-2 infection incidence rates (IR), and their 95% confidence intervals (CI), per 100,000 person-days during five time-period: days <7 days, 8–14 days, 15–21 days, 22–28 days, and >28 days from receipt of the first BNT162b2 dose. Individuals stopped contributing person-days once SARS-CoV-2 infection was confirmed or at the study end date, whichever came first. We used Poisson regression to calculate incidence rate ratios (IRR) and their 95% CI for the latter four time-periods relative to IR during the first seven days from the first BNT162b2 dose. Statistical analyses were performed using Stata Statistical Software, Release 16.1 (StataCorp., College Station, Texas).

## Results

The included 199,219 individuals contributed 6,521,124 person-days of follow up. SARS-CoV-2 infection was confirmed in 1,877 (0.9%), of which 365 (19.5%) occurred after receipt of a second BNT162b2 dose. The median time from first vaccination to SARS-CoV-2 confirmation was 11.9 days (IQR 7.7–18.2). Compared with those without SARS-CoV-2 infection, infected individuals were significantly older, and more likely to have co-existing medical conditions (Table). Cough (1,018, 54.2%) and fever (745, 39.7%) were the most frequent presenting symptoms, while infections were asymptomatic in 489 (26.1%). High-flow nasal oxygen or non-invasive ventilation was required for 28 (1.5%) individuals, invasive mechanical ventilation for 11 (0.6%), and oxygen via face mask or nasal cannula for 84 (4.5%). Five individuals (median age 81 years, range 52–93) had fatal COVID-19, all with multiple comorbidities, and most with infected household contacts. For fatal COVID-19 cases, symptoms started after a median of 12 days (range 7–25) after the first BNT162b2 dose.

**Table.** Baseline characteristics of the study cohort.

|  | **Total study**<br>**cohort**<br>**(n = 199,219)** | **Group without SARS-**<br>**CoV-2 infection**<br>**(n = 197,342 )** | **Group with SARS-**<br>**CoV-2 infection**<br>**(n = 1,877)** | **P value** |
| --- | --- | --- | --- | --- |
| BNT162b2 doses received |  |  |  |  |
| One dose | 129,462 (65%) | 129,950 (65%) | 1512 (80.5%) |  |
| Two doses | 69,757 (35%) | 69,392 (35%) | 365 (19.5%) |  |
| Female sex | 83,034 (41.7%) | 82,214 (41.7%) | 820 (43.7%) | 0.076 |
| Age (years) | 42 (33–55) | 42 (33–55) | 44 (35–54) | <0.001 |
| Age groups (years) |  |  |  |  |
| 16–24 | 13,826 ( 6.9%) | 13,764 (7%) | 62 (3.3%) |  |
| 25–34 | 43,302 (21.7%) | 42,934 (21.8%) | 368 (19.6%) |  |
| 35–44 | 51,934 (26.1%) | 51,384 (26%) | 550 (29.3%) |  |
| 45–54 | 39,274 (19.7%) | 38,823 (19.7%) | 451 (24%) |  |
| 55–64 | 31,257 (15.7%) | 30,978 (15.7%) | 279 (14.9%) |  |
| ≥65 | 19,626 (9.9%) | 19,459 (9.9%) | 167 (8.9%) |  |
| Co-existing medical conditions |  |  |  |  |
| Diabetes Mellitus | 39,994 (20.1%) | 39,550 (20%) | 444 (23.7%) | <0.001 |
| Hypertension | 42,994 (21.6%) | 42,512 (21.5%) | 482 (25.7%) | <0.001 |
| Chronic heart disease | 10,308 (5.2%) | 10,190 (5.2%) | 118 (6.3%) | <0.029 |
| Chronic lung disease | 19,477 (9.8%) | 19,270 (9.8%) | 207 (11%) | 0.067 |
| Chronic kidney disease | 4,272 (2.1%) | 4,209 (2.13%) | 63 (3.4%) | <0.001 |
| Chronic liver disease | 1,781 (0.9%) | 1,754 (0.9%) | 27 (1.4%) | 0.012 |
| Malignant disease | 3,724 (1.9%) | 3,691 (1.9%) | 33 (1.8%) | 0.72 |
| Organ transplant recipient | 653 ( 0.33%) | 640 ( 0.32%) | 13 ( 0.69%) | 0.005 |

Data are presented as number (%) or median (interquartile range). P values were derived from Pearson's chi-
squared or Wilcoxon rank-sum test, as appropriate.

SARS-CoV-2 IR was 28.63/100,000 person-days (95% CI 25.96–31.58) during the first seven days post-vaccination, and 27.19/100,000 person-days (95% CI 25.32–29.2) during days 8–14. Compared with the first 7-day post-vaccination period, the IR was significantly lower during days 15–21 (IRR 0.342, 95% CI 0.297–0.394, P <0.001), days 22–28 (IRR 0.171, 95% CI 0.142–0.205, P <0.001), and >28 days (IRR 0.153, 95% CI 0.129–0.182, P <0.001). Similarly, in comparison with the first 7-day post-vaccination period, the IRR for severe COVID-19 decreased significantly during days 15–21 (IRR 0.243, 95% CI 0.136– 0.434, P <0.001), days 22–28 (IRR 0.171, 95% CI 0.087–0.335, P <0.001), and >28 days (IRR 0.067, 95% CI 00.028–0.161, P <0.001) (Figure).

**Figure.**
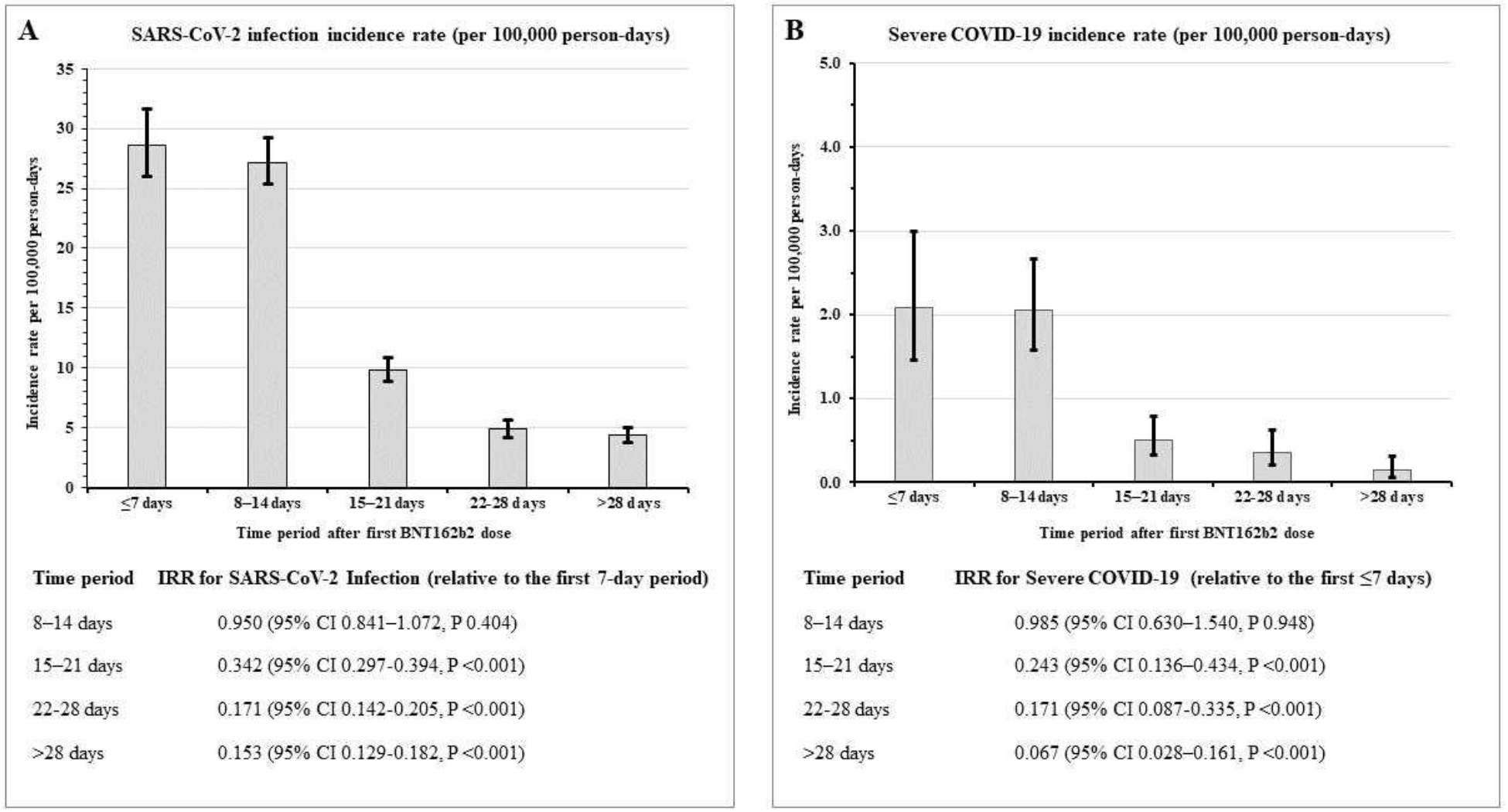
SARS-CoV-2 incidence rate by time-period following the first dose of BNT162b2 vaccination. Panel A, SARS-CoV-2 infection incidence rate (per 100,000 person-days), Panel B, Severe COVID-19 incidence rate (per 100,000 person-days). COVID-19, Coronavirus Disease 2019; CI, confidence interval; IRR, incidence rate ratio; SARS-CoV-2, Severe Acute Respiratory Syndrome Coronavirus 2. I bars indicate 95% confidence intervals.

Our findings are consistent with those shown previously in a community setting and in healthcare workers, but our report is the first to include an entire national cohort of BNT162b2 recipients.[2, 4] We also demonstrated a significant reduction in risk of severe COVID-19. This is particularly important, given its potential to reduce COVID-19-associated morbidity and mortality, and decrease its impact on healthcare resource utilization.[5]

We found relatively high SARS-CoV-2 IR during the first two weeks following receipt of the first BNT162b2 dose. While the vaccine’s protective effect may not be apparent during the first two weeks after BNT162b2 vaccination,[1] recipients may wrongly perceive themselves to be at a reduced risk of SARS-CoV-2 infection and become less adherent to nonpharmacological preventive measures such as social distancing and face covering.[6] Careful education and counselling during the vaccination process could help efforts to minimise such risk compensation behaviour.

SARS-CoV-2 variants of concerns (VOC) such B.1.1.7 lineage (first reported in the United Kingdom), and B.1.351 lineage (first reported in South Africa), are known to have been circulating in Qatar during the study’s follow up period.[7] There has been concern that VOC may reduce the effectiveness of some COVID-19 vaccines.[8] However, BNT162b2 elicits high neutralising antibodies titres against B.1.1.7 and B.1.351 lineages.[9] Moreover, recently announced results from an ongoing phase 3 BNT162b2 randomised trial suggest it is highly effectiveness against B.1.351 lineage.[10]

Deaths in our study mostly occurred in older individuals with multiple co-morbidities. Vaccine effectiveness is generally lower in such groups.[2] Notably, infected household contacts were identified in three out of the five fatal COVID-19 cases, reinforcing the importance of wide vaccine rollout to maximise protection of the most vulnerable.[5]

The limitations of this study include its observational nature and the lack of a non-vaccinated control group. We used the first 7-day post-vaccination period, during which no vaccine effectiveness is expected, as a reference to assess the vaccine’s protective benefits in later time-periods. Overall, our results are consistent with an early protective effect of BNT162b2 against all degrees of SARS-CoV-2 severity. It is anticipated that, in addition to ongoing nonpharmacological interventions, broader vaccine coverage will contribute to the national and global pandemic control efforts.

## Ethical issues

The study was approved by Hamad Medical Corporation’s Institutional Review Board with a waiver of informed consent (MRC-01-21-207).

## Data Availability

The datasets generated and/or analyzed during the current study are available from the corresponding author on reasonable request.

## Conflict of interests

The authors declare no conflict of interests in relation to this manuscript.

## Funding

The publication of this report was funded by Qatar National Library. No other funding was required.

## Acknowledgments

We would like to thank Hussam Alsoub and Faraj S. Howady for their support during the preparation of this report.

## Notes

### Competing Interest Statement

The authors have declared no competing interest.

### Author Declarations

The study was approved by Hamad Medical Corporation Institutional Review Board with a waiver of informed consent (MRC-01-21-207).

